# Long Covid in adults discharged from UK hospitals after Covid-19: A prospective, multicentre cohort study using the ISARIC WHO Clinical Characterisation Protocol

**DOI:** 10.1101/2021.03.18.21253888

**Authors:** Louise Sigfrid, Thomas M Drake, Ellen Pauley, Edwin C Jesudason, Piero Olliaro, Wei Shen Lim, Annelise Gillesen, Colin Berry, David J Lowe, Joanne McPeake, Nazir Lone, Muge Cevik, Daniel J Munblit, Anna Casey, Peter Bannister, Clark D Russell, Lynsey Goodwin, Antonia Ho, Lance Turtle, Margaret E O’Hara, Claire Hastie, Chloe Donohue, Rebecca Spencer, Janet Harrison, Cara Donegan, Alison Gummery, Hayley Hardwick, Claire E Hastie, Laura Merson, Gail Carson, J Kenneth Baillie, Peter JM Openshaw, Ewen M Harrison, Annemarie B Docherty, Malcom G Semple, Janet T Scott, ISARIC global follow-up working group, ISARIC4C investigators

## Abstract

**Evidence before this study:** It is emerging that long-term symptoms are often present in people who have had acute COVID-19 disease. These symptoms affect a range of organ systems including respiratory, cardiovascular and neurological systems. It is not clear how many patients who required hospitalisation develop these symptoms and the impact they have on quality of life. We searched PubMed on 24^th^ March 2021 using the terms ‘COVID-19’, ‘long-Covid’, ‘long-term’ and ‘outcomes’. This was supplemented by hand searching relevant references and news reports. We identified several small studies focussing on specific symptoms or diseases, studies of patients in community settings and studies of patients who were hospitalised for acute COVID-19 in Italy, Russia and China. There were no peer-reviewed published data at the time of searching which captured outcomes of patients within the United Kingdom.

**Added Value of this study:** We found that over half of patients reported not feeling fully recovered several months after onset of Covid-19 symptoms. The most common symptoms reported were fatigue, followed by breathlessness. Patients reported significant increases in new or worse disability, increases in MRC dyspnoea scale and worse quality of life as measured by EQ5D-5L summary index at the time of follow-up compared to before onset of acute COVID-19 symptoms. Our study found that women, in particular women under the age of 50 were significantly more likely to not feel fully recovered, were more breathless, more fatigued and had higher rates of new or worsened disability, even after taking severity of acute disease into account when compared to men of the same age.

**Implications of all available evidence:** Long-term symptoms following hospitalisation for COVID-19 are very common and have significant impacts on quality of life. Women under 50 were most likely to have the worst outcomes. Policy makers need to ensure there is long-term support for people experiencing long-Covid and should plan for lasting long-term population morbidity. Funding for research to understand mechanisms underlying long-Covid and identify potential interventions for testing in randomised trials is urgently required.

**Structured Abstract:** *Background:* This study sought to establish the long-term effects of Covid-19 following hospitalisation.

*Methods:* 327 hospitalised participants, with SARS-CoV-2 infection were recruited into a prospective multicentre cohort study at least 3 months post-discharge. The primary outcome was self-reported recovery at least ninety days after initial Covid-19 symptom onset. Secondary outcomes included new symptoms, disability (Washington group short scale), breathlessness (MRC Dyspnoea scale) and quality of life (EQ5D-5L).

*Findings:* 55% of participants reported not feeling fully recovered. 93% reported persistent symptoms, with fatigue the most common (83%), followed by breathlessness (54%). 47% reported an increase in MRC dyspnoea scale of at least one grade. New or worse disability was reported by 24% of participants. The EQ5D-5L summary index was significantly worse at follow-up (median difference 0.1 points on a scale of 0 to 1, IQR: −0.2 to 0.0). Females under the age of 50 years were five times less likely to report feeling recovered (adjusted OR 5.09, 95% CI 1.64 to 15.74), were more likely to have greater disability (adjusted OR 4.22, 95% CI 1.12 to 15.94), twice as likely to report worse fatigue (adjusted OR 2.06, 95% CI 0.81 to 3.31) and seven times more likely to become more breathless (adjusted OR 7.15, 95% CI 2.24 to 22.83) than men of the same age.

*Interpretation:* Survivors of Covid-19 experienced long-term symptoms, new disability, increased breathlessness, and reduced quality of life. These findings were present in young, previously healthy working age adults, and were most common in younger females. Role of the funder: The study sponsors and funders had no role in the study design, collection, analysis, interpretation of data, writing of the report, or the decision to submit the article for publication. Investigators were independent from funders and the authors have full access to all of the data, including any statistical analysis and tables.

## Introduction

Our understanding of long-term outcomes after acute Covid-19 disease remains limited. It is becoming increasingly evident that some patients who have had acute Covid-19 go on to experience persistent symptoms, known as long-Covid or post-covid syndrome^1^. Several studies in hospitalised and community settings have identified that those with Covid-19 frequently develop long-term symptoms and a range of sequelae affecting the kidneys, lungs and heart^1–5^. These symptoms appear to overlap with other post-viral syndromes and with the challenges faced by patients recovering from other critical illness with post-intensive care syndrome (PICS), such as muscle weakness, fatigue, and sleep disturbance ^6–10^. Yet, understanding the impact Covid-19 has on patient reported outcome measures, including quality of life, has not yet been fully characterised ^11^.

Many clinical trials or studies that aim to characterise the immediate course of Covid-19 have used mortality as a primary outcome^12,13^. This has demonstrated that patients in older age groups and those who have pre-existing comorbidities are at higher risk of dying from the disease^14–17^. Nonetheless, most people with Covid-19 will survive the initial acute infection and data on what happens to these individuals in the long-term are lacking. The large number of people affected by Covid-19 and the growing evidence of long-term sequelae highlights the importance for policy makers, society and healthcare systems to understand the difficulties faced by those suffering from long-Covid^4,18,19^. Understanding the burden of disease, and who is at greatest risk of developing long-term complications, may help to target preventative strategies and provide effective support for affected individuals to improve Covid-19 outcomes and reduce risk of widening health inequalities by inadequate rehabilitation and recovery support. Identifying which patient groups are most likely to be affected could provide data to guide policy and aid future research to identify disease mechanisms, formulate and test new interventions.

The objective of this study is to characterise long-term patient reported outcomes in individuals who survived hospitalisation for Covid-19, in those who engaged with follow-up, using the International Severe Acute Respiratory and emerging Infections Consortium (ISARIC) WHO Clinical Characterisation Protocol (CCP-UK) and follow-up protocol ^20^.

## Methods

### Study design and setting

The ISARIC WHO CCP protocol was first developed by international consensus in 2012 to respond to any emerging or re-emerging pathogen of public health interest^21^. It was activated in the UK in response to the SARS-CoV-2 pandemic on 17th January 2020. Study information including the CCP-UK and follow-up protocol, standardised case report forms, study information and consent forms, are available on the <u>ISARIC4C.net</u> website. Hospitals providing acute care throughout the United Kingdom are eligible to enrol participants into the study. This analysis is reported in line with the Strengthening the Reporting of Observational Studies in Epidemiology (STROBE) guidelines^22^.

### Participants

Patients aged 18 years and over, admitted to hospital between 17^th^ January to 5^th^ October 2020 with confirmed or highly suspected SARS-CoV-2 infection at 31 centres, who consented to be contacted for follow-up and were discharged at least 90 days ago were eligible for inclusion. Confirmation of SARS-CoV-2 was by reverse-transcriptase polymerase chain reaction (RT-PCR). Individuals with highly suspected, Covid-19 were also eligible for inclusion, given that SARS-CoV-2 was an emergent pathogen in the earlier stages of the pandemic and laboratory confirmation was dependent on local availability of PCR testing.

### Variables

Patient questionnaires for adults were developed by a multidisciplinary team of researchers, clinicians and psychologists through a series of meetings and e-mail iterations^21^. These were piloted in three countries before being finalised. The UK version was piloted with patients at sites in Liverpool and Glasgow. Patient questionnaires were designed to allow self-assessment via post, or clinician led follow up via telephone, or in outpatient clinic, to support wide dissemination. All surviving patients who consented to be contacted following discharge and for which a valid address or phone number were provided were contacted. Questionnaires were posted from the Outbreak Laboratory coordinating centre at the University of Liverpool, UK, with a prepaid, self-addressed envelope for returning the questionnaire. A combination of postal and telephone follow-up was used to improve response rates. Those who did not respond by post and who had a valid phone number were followed up by telephone or in outpatient clinic by local study investigators. Participants completed one questionnaire as part of this study, so there were no repeat measures. Data from responses were entered onto a Research Electronic Capture (REDCap) Database system hosted at the University of Oxford and linked with data documented during the admission with acute Covid-19 for the analysis.

Explanatory variables at the time of hospital admission, including age, sex, pre-existing comorbidities, and treatment received during the hospital admission were recorded. Maximum severity of Covid-19 during the acute hospital admission with Covid-19 was classified using the WHO COVID-19 ordinal severity scale^23^. This scale comprised of 4 levels of severity which were relevant to our in-hospital cohort; level 3 - did not receive supplemental oxygen, level 4 - received supplemental oxygen, level 5 - received high flow oxygen or NIV non-invasive ventilation (HFNC, NIV), and levels 6 and 7 - received invasive mechanical ventilation or admission to critical care)^23^. We also used the WHO severity scale to account for in-hospital severity in our modelling approach^23,24^.

### Outcomes

The primary outcome was self-reported recovery at 3 to 12 months following initial Covid-19 symptoms. Secondary outcomes included persistent or new symptoms, new or worsened disability assessed using the Washington Disability Group (WG) Short Form^25^, breathlessness measured using the Medical Research Council (MRC) dyspnoea scale^26^, fatigue measured on a 1 to 10 visual analogue scale (VAS) where zero is no fatigue and ten is worst possible fatigue, and quality of life using the EuroQol^®^ EQ5D-5L instrument (supplementary appendix 2)^27^. The MRC dyspnoea scale was developed to grade the effect of breathlessness on daily activities^26^. This 5-point scale measures perceived respiratory disability, with 1 being no breathlessness and 5 being unable to undertake activities of daily living due to breathlessness^26^. The WG Short Set tool includes six questions on functioning (vision, hearing, mobility, cognition, self-care, communication)^25^. These questions reflect a bio-psychosocial model of disability by describing level of disability and probe aspects of disability which may limit an individual’s participation in society. This tool has been shown to detect the majority of disabilities and is standardised for use globally^25^. The EuroQol^®^EQ5D-5L tool was used to measure psychosocial health and quality of life^27^. The tool covers five dimensions: mobility, self-care, usual activities, pain/discomfort and anxiety/depression. The person indicates his/her health state for each of the five dimensions. To compare the change in EQ5D-5L at the time of follow-up to before Covid-19 onset, we asked patients the same questions contained in the EQ5D-5L with the tense altered to ask specifically about pre-Covid-19 state.

### Statistical methods

Categorical data were summarised as frequencies and percentages, and continuous data as median, alongside the corresponding interquartile range (IQR) presented as the 25^th^ and 75^th^ centile values. To test for differences across comparison groups in categorical data, we used Fisher’s exact test and for continuous data, used the Wilcoxon rank-sum test for two-sample testing and Kruskal-Wallis where there were more than 2 groups.

For disability, breathlessness, and EQ5D-5L index (health state), we calculated the change in value reported by participants before onset of their Covid-19 illness compared to the follow up assessment. For health state at the follow up assessment, we used the EQ5D-5L with the English standardised valuation study protocol (EQ-VT) value set, developed by the EuroQol group on the composite time trade-off (cTTO) valuation^27^. Overall changes in summary health index, before and after Covid-19 onset, were summarised for the cohort using the Paretian Classification of Health Change (PCHC) method.^28,29^ Summary EQ5D-5L indices and change in summary EQ5D-5L index were measured on a scale of 0 to 1, with 1 being perfect health and 0 being worst health imaginable. We calculated both the overall estimates and estimates for individual EQ5D-5L dimensions.

We created models to adjust for age, sex, presence of comorbidities and in-hospital severity of Covid-19, according to the maximum level of respiratory support that was required. Multilevel logistic regression was used for binary outcomes, and linear regression models were used for continuous outcomes. In both model types, we adjusted for the effects of explanatory variables using fixed-effects and centre by including a random-effects term. Final model selection was guided by minimisation of the Akaike information criterion (AIC). Variables were only included in the model if they were present during the first hospital admission for Covid-19. All models were checked for first order interactions and any meaningful interactions were retained and incorporated as dummy variables. Effect estimates are presented as odds ratios for binary outcomes or mean differences for continuous outcomes, alongside the corresponding 95% confidence interval (95% CI). Statistical analyses were performed using R version 3.6.3 (R Foundation for Statistical Computing, Vienna, AUT) with the tidyverse, finalfit, eq5d and Hmisc packages. Statistical significance was taken at the level of P ≤0.05.

### Public and patient involvement

This was an urgent public health research study in response to a public health emergency of international concern. Patients and the public were therefore not involved in the design, of the acute phase rapid response research. However, patients and people living with long covid were involved in the design, conduct and interpretation of the follow up study. The follow up data collection survey and associated patient information was informed by the founding members of the Long Covid support group, who themselves are living with long Covid. The survey was also piloted in several settings in the UK with patients affected by Covid-19 from different demographics, and feedback incorporated into the final version. This included suggestions on the data on symptoms collected and the way questions were asked as well as on the patient information. The results and interpretation of the findings and final manuscript were informed by members of the Long Covid support group.

## Results

Of the 2150 eligible people in the CCP-UK study who were discharged from their acute admission alive, 40.1% (862/2150) provided consent to be contacted for follow-up. Of these, 97.8% (843/862) were contacted. From these 843 people, 97.8% (824/843) were 18 or over and 53.7% (443/824) completed the follow-up questionnaire. Finally, of respondents 73.8% (327/443) responded 90 days or more after symptom onset. Included participants completed the follow-up questionnaire through self-assessment (71 6% 234/327), telephone (24 5% 80/327) or in outpatient clinic (4 0% 13/327, figure 1). The median follow-up time from symptom onset was 222 days (IQR: 189 to 269 days, range: 112 to 343 days, table 1).

**Figure 1.**
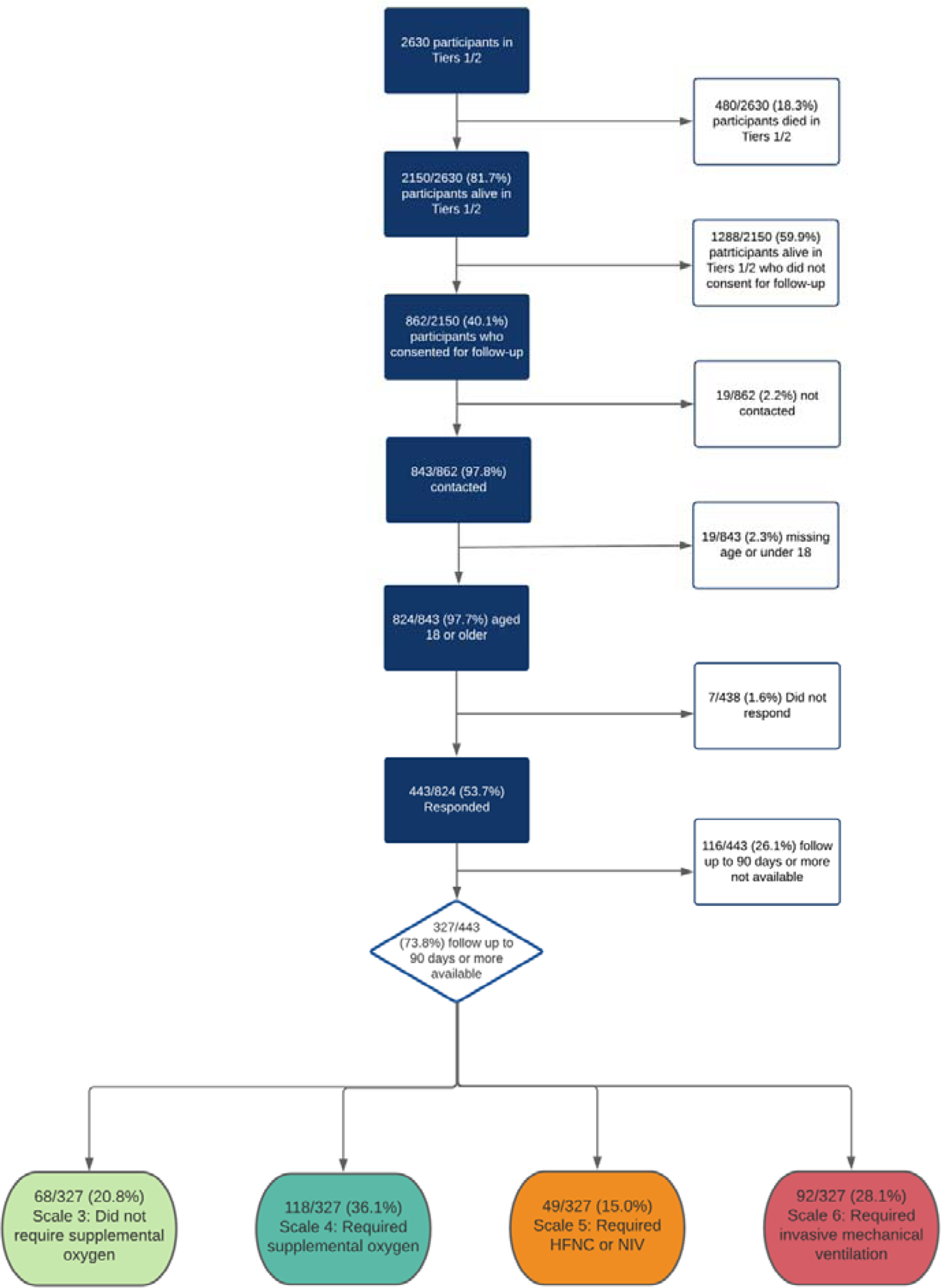
Patient inclusion flowchart

**Table 1.** Characteristics of participants who responded severity)

|  |  | Recovered or unsure | Does not feel fully recovered | (Missing) | p-value |
| --- | --- | --- | --- | --- | --- |
| Total N (%) |  | 144 (44.0) | 179 (54.7) | 4 (1.2) |  |
| Age | Median (IQR) | 60.5 (53.2 to 69.8) | 59.4 (50.3 to 66.7) | 54.2 (48.2 to 58.5) | 0.089 |
|  | Under 50 | 26 (37.1) | 43 (61.4) | 1 (1.4) |  |
|  | 50 to 69 | 83 (42.6) | 109 (55.9) | 3 (1.5) |  |
|  | Over 70 | 35 (56.5) | 27 (43.5) | 0 (0.0) |  |
| Sex at Birth | Male | 87 (45.3) | 103 (53.6) | 2 (1.0) | 0.683 |
|  | Female | 57 (42.2) | 76 (56.3) | 2 (1.5) |  |
| Ethnicity | White | 115 (43.4) | 147 (55.5) | 3 (1.1) | 0.206 |
|  | South Asian | 2 (25.0) | 6 (75.0) | 0 (0.0) |  |
|  | East Asian | 3 (75.0) | 1 (25.0) | 0 (0.0) |  |
|  | Black | 10 (66.7) | 5 (33.3) | 0 (0.0) |  |
|  | Other Ethnic Minority | 10 (47.6) | 10 (47.6) | 1 (4.8) |  |
|  | (Missing) | 4 (28.6) | 10 (71.4) | 0 (0.0) |  |
| Smoking | Never Smoked | 84 (47.7) | 90 (51.1) | 2 (1.1) | 0.892 |
|  | Current Smoker | 4 (57.1) | 3 (42.9) | 0 (0.0) |  |
|  | Former Smoker | 43 (46.7) | 47 (51.1) | 2 (2.2) |  |
|  | (Missing) | 13 (25.0) | 39 (75.0) | 0 (0.0) |  |
| Diabetes | No | 112 (44.1) | 138 (54.3) | 4 (1.6) | 0.972 |
|  | Yes | 27 (43.5) | 35 (56.5) |  |  |
| Obesity (as defined by clinical staff) | No | 116 (45.7) | 136 (53.5) | 2 (0.8) | 0.291 |
|  | Yes | 20 (35.7) | 34 (60.7) | 2 (3.6) |  |
|  | (Missing) | 8 (47.1) | 9 (52.9) | 0 (0.0) |  |
| Chronic cardiac disease | No | 119 (43.8) | 149 (54.8) | 4 (1.5) | 1.000 |
|  | Yes | 20 (45.5) | 24 (54.5) | 0 (0.0) |  |
|  | (Missing) | 5 (45.5) | 6 (54.5) | 0 (0.0) |  |
| Chronic pulmonary disease (not asthma) | No | 126 (43.4) | 160 (55.2) | 4 (1.4) | 0.592 |
|  | Yes | 12 (52.2) | 11 (47.8) | 0 (0.0) |  |
|  | (Missing) | 6 (42.9) | 8 (57.1) | 0 (0.0) |  |
| Asthma (physician diagnosed) | No | 114 (45.2) | 135 (53.6) | 3 (1.2) | 0.410 |
|  | Yes | 25 (38.5) | 39 (60.0) | 1 (1.5) |  |
|  | (Missing) | 5 (50.0) | 5 (50.0) | 0 (0.0) |  |
| Chronic kidney disease | No | 131 (44.0) | 163 (54.7) | 4 (1.3) | 1.000 |
|  | Yes | 8 (44.4) | 10 (55.6) | 0 (0.0) |  |
|  | (Missing) | 5 (45.5) | 6 (54.5) | 0 (0.0) |  |
| Malignant neoplasm | No | 136 (44.4) | 166 (54.2) | 4 (1.3) | 1.000 |
|  | Yes | 4 (40.0) | 6 (60.0) | 0 (0.0) |  |
|  | (Missing) | 4 (36.4) | 7 (63.6) | 0 (0.0) |  |
| Rheumatologic disorder | No | 131 (45.2) | 155 (53.4) | 4 (1.4) | 0.334 |
|  | Yes | 8 (33.3) | 16 (66.7) | 0 (0.0) |  |
|  | (Missing) | 5 (38.5) | 8 (61.5) | 0 (0.0) |  |
| ISARIC4C Mortality Score (predicted severity) | Median (IQR) | 7.0 (5.0 to 9.0) | 6.0 (4.0 to 9.0) | 5.5 (5.0 to 6.0) | 0.648 |
| Severity | Scale 3 (did not receive supplemental oxygen) | 34 (50.0) | 33 (48.5) | 1 (1.5) | 0.001 |
|  | Scale 4 (received supplemental oxygen) | 63 (53.4) | 53 (44.9) | 2 (1.7) |  |
|  | Scale 5 (received HFNC or NIV) | 22 (44.9) | 27 (55.1) | 0 (0.0) |  |
|  | Scale 6 or 7 (received invasive mechanical ventilation or critical care) | 25 (27.2) | 66 (71.7) | 1 (1.1) |  |
| Critical care admission | Ward level care only | 99 (50.3) | 96 (48.7) | 2 (1.0) | 0.008 |
|  | Admitted to Critical Care | 45 (34.6) | 83 (63.8) | 2 (1.5) |  |
| Length of stay (days) | Median (IQR) | 8.0 (5.0 to 13.0) | 11.0 (6.2 to 25.0) | 4.5 (3.0 to 7.8) | <0.001 |
| Time from symptoms to completing survey (days) | Median (IQR) | 221.0 (190.0 to 245.0) | 224.0 (188.0 to 292.0) | 210.0 (194.2 to 218.5) | 0.419 |
| Time from discharge to completing survey (days) | Median (IQR) | 200.0 (177.0 to 230.0) | 199.0 (161.0 to 268.0) | 195.0 (178.0 to 208.2) | 0.846 |

HFNC – High flow nasal cannulae, NIV – Noninvasive ventilation, IQR – Interquartile range, presented as 25<sup>th</sup> to 75<sup>th</sup> centiles. Numbers are presented as N (%), unless otherwise denoted as a continuous variable.

### Participant characteristics

Table 1 shows the characteristics of participants who responded. The majority of participants were male (58 7%, 192/327), with a median age of 59 7 (25^th^ centile 51.7 to 75^th^ centile 67 7) years and of white ethnicity (81 0%, 265/327, table 1). Asthma (19 9%, 65/327) and diabetes (19 0%, 62/327) were the most common comorbidities (table 1). Compared with the study population who were contacted and did not respond, respondents were significantly more likely to be of white ethnicity (81 0%, 265/327 participants versus 66 6%, 331/544 of non-respondents), were more likely to be ex-smokers (28 1%, 92/327 participants versus 24 7% 123/497 of non-respondents) and were more likely to have been admitted to critical care (39 8% 130/327 in participants versus 26 8% 133/497 in non-respondents, supplementary table 1).

### Outcomes and symptoms

Of 327 participants, 54 7% (179/327) did not feel they had fully recovered at the time of follow-up. At the univariable level, there were no associations between not feeling recovered and the risk factors of age, sex, ethnicity, and comorbidities (table 1) but we found patients with a higher severity of acute disease were significantly more likely not to feel recovered. Persistent or new symptoms were reported by 93 3% (305/327) participants (table 2). The most frequently reported symptoms were fatigue 82 8% (255/308), shortness of breath 53 5% (175/327), and problems sleeping 46 2% 151/327, figure 2A).

**Table 2.**
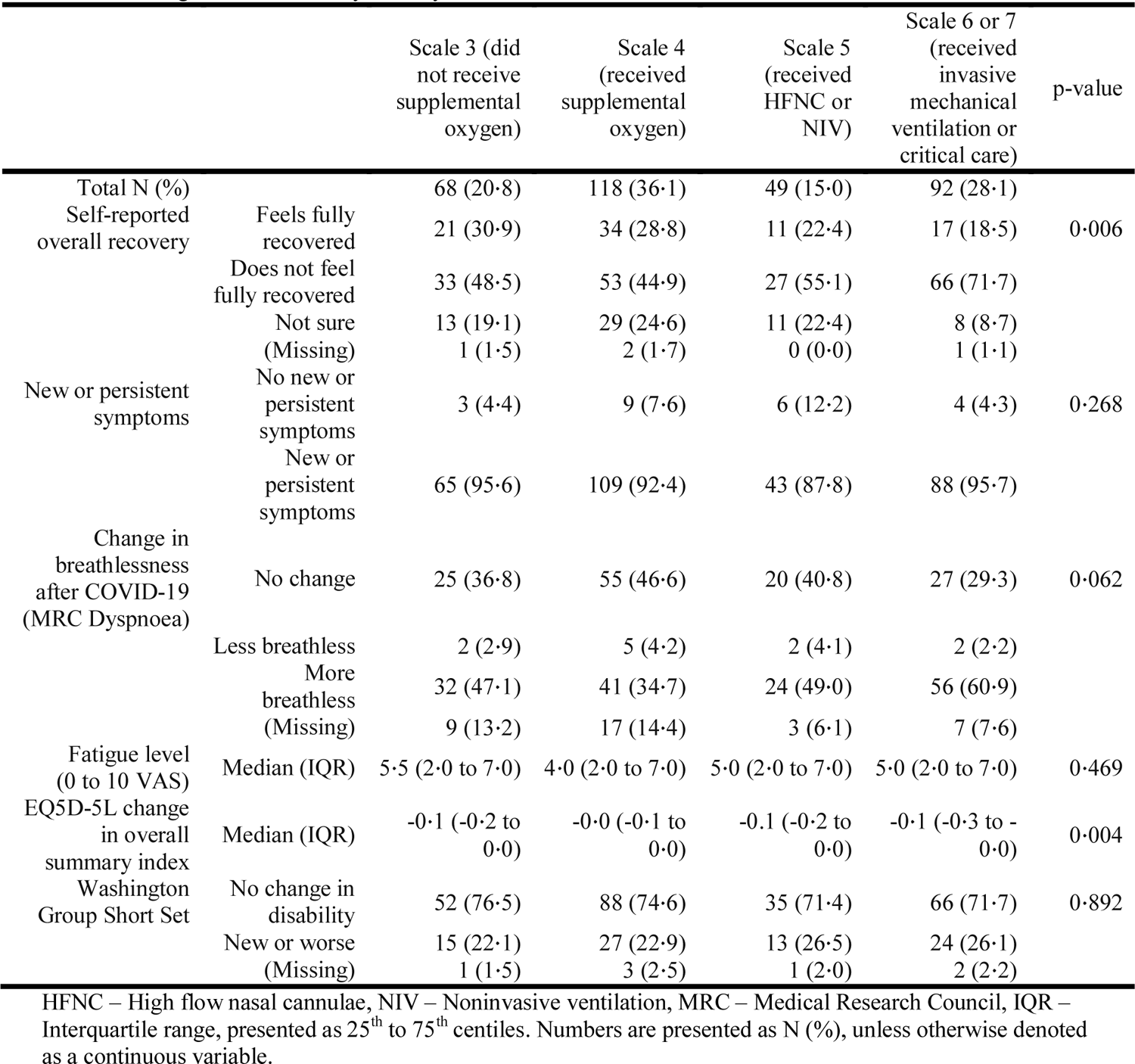
Long-term outcomes by severity of acute Covid-19.

**Figure 2.**
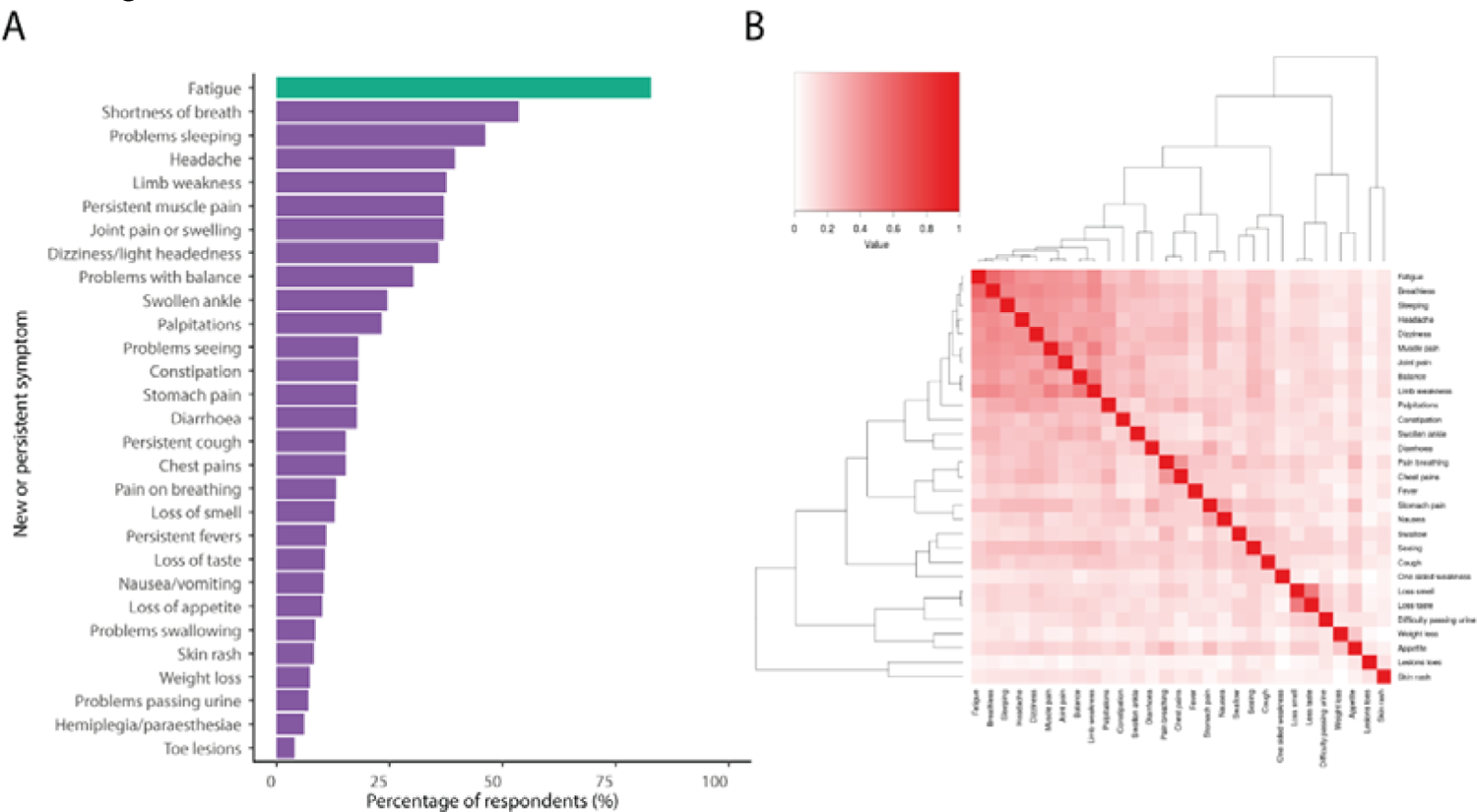
Proportion of new or persistent symptoms occurring (figure 2A) and their co-occurrence with each other (figure 2B). For figure 2A, fatigue is coloured in green as this outcome was derived from the fatigue visual analogue outcome, where a fatigue rating of 2 or greater was considered as the presence of the fatigue symptom (see supplementary table 7 for raw values). Erectile dysfunction affected 23 4% (45/192) of males included, not shown as figure 2A presents data for any sex. For figure 2B, the Jaccard similarity index was calculated and presented as intensity of red colour, with 0 (white) being no co-occurrence and 1 (bright red) being always co-occurring.

A heatmap and dendrogram of symptom co-occurrence identified two major clusters of symptoms (figure 2B); a fatigue, myalgia and sensorineural deficits cluster and an olfactory, appetite and urinary cluster (loss of smell, loss of taste, difficulty passing urine, weight loss and loss of appetite). Within the fatigue, myalgia and sensorineural deficits cluster, there was a distinct minor cluster affecting movement (muscle pain, joint pain, balance and limb weakness).

In addition to symptomatic breathlessness, 46 8% (153/327) of participants reported increased breathlessness compared to their pre-Covid-19 baseline. Overall, change in breathlessness was not affected by age or number of comorbidities (figure 3), but was significantly higher in females compared to males (41 7%, 80/192 in males versus 54 1% 73/135 in females). Of participants with a pre-Covid-19 MRC grade 1, 34 0% (73/215) reported an increase to grade 2, and 25 6% (55/213) reported an increase to grades 3-5 at time of follow-up (figure 4A to 4C). Proportionally, those who were admitted to critical care were more likely to have a higher MRC dyspnoea grade at the time of follow-up.

**Figure 3.**
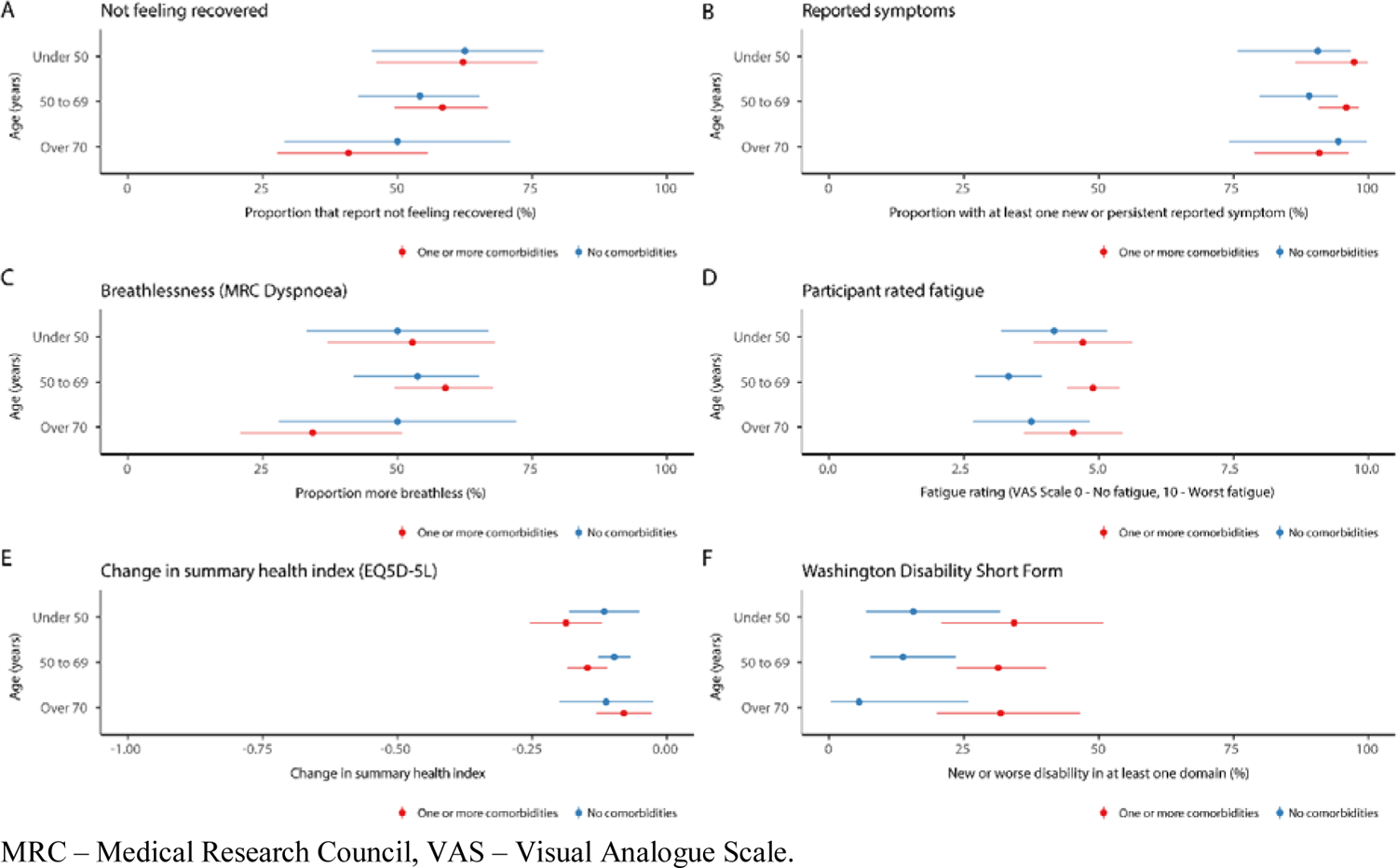
Outcomes stratified by age and presence of one or more comorbidities. Figure 3A – Proportion of participants not feeling fully recovered; Figure 3B – Proportion of participants with new or persistent symptoms; Figure 3C - Proportion of participants with increased breathlessness as measured by MRC dyspnoea scale; Figure 3D – Participant rated fatigue on 0 to 10 VAS; Figure 3E – Change in overall EQ5D-5L summary health index; Figure 3F – presence of new or worse disability in at least one Washington Group disability domain.

**Figure 4.**
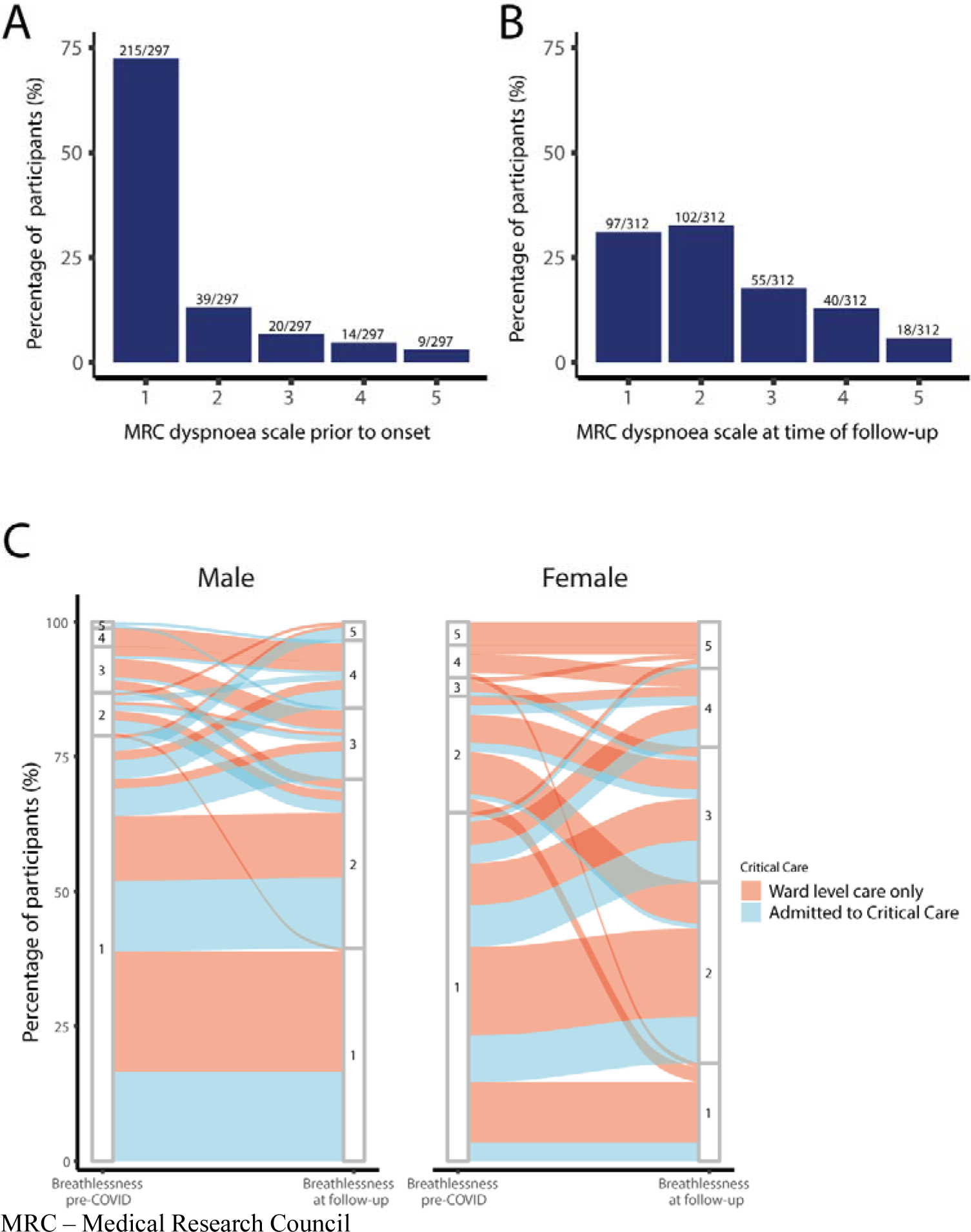
MRC Dyspnoea scale prior to Covid-19 onset and at the time of follow-up. Figure 4A – MRC dyspnoea scale reported prior to onset of Covid-19 symptoms; Figure 4B – MRC dyspnoea scale at the time of follow-up; Figure 4C – Alluvial plot of proportion of the changes in proportion of men and women in each MRC scale grade before symptom onset and at time of follow-up, stratified in each sex group by admission to critical care. In figure 4C, for females, there are greater numbers of participants who begin at MRC 1 and transition to higher levels on the scale compared with males.

Overall, intensity of fatigue was unrelated to age or disease severity in hospital (figure 3, table 2), but females were found to have significantly increased levels of fatigue compared with males (median fatigue 0-10 VAS score, males 4 0, IQR 2.0 to 6; versus females 6 0, IQR 2 0 to 7 0, supplementary table 2, supplementary figure 1).

New or worsened disability in at least one Washington Group domain was experienced by 24 2% (79/327). This did not change by in-hospital Covid-19 severity (table 2) or comorbidities (figure 3). Females reported a greater number of new or worsened disabilities compared to males (20 3%, 39/192 in males compared with 29 6%, 40/135 in females, supplementary table 2). The most affected domain was walking and mobility (33 3% 109/327 new mild or worsened disability, 6 4% 21/327 new moderate or worsened disability and 0 3% 1/327 new severe or worsened disability, supplementary table 3), followed by memory and concentration (30 0% 90/327 new mild or worsened disability, 9.8% 32/327 new moderate or worsened disability). There were significant differences in domains affected by sex, with females reporting significantly higher levels of visual disabilities (12 0% 23/192 new mild or worsened disability for males versus 25 2% 34/135 new mild or worsened disability for females, supplementary table 3), higher levels of walking disability (28.6% 55/192 new mild or worsened disability for males versus 40 0% 54/135 new or worsened mild disability for females; 5 6% 11/192 new moderate or worsened disability for males versus 7 4% 10/327 new moderate or worsened disability for females) and memory disability (27 1% 52/192 new mild or worsened disability for males versus 34 1% 46/135 new mild or worsened disability for females; 7 3% 14/192 new moderate or worsened disability for males versus 13 3% 18/135 new moderate or worsened disability for females, supplementary table 4).

**Table 3.**
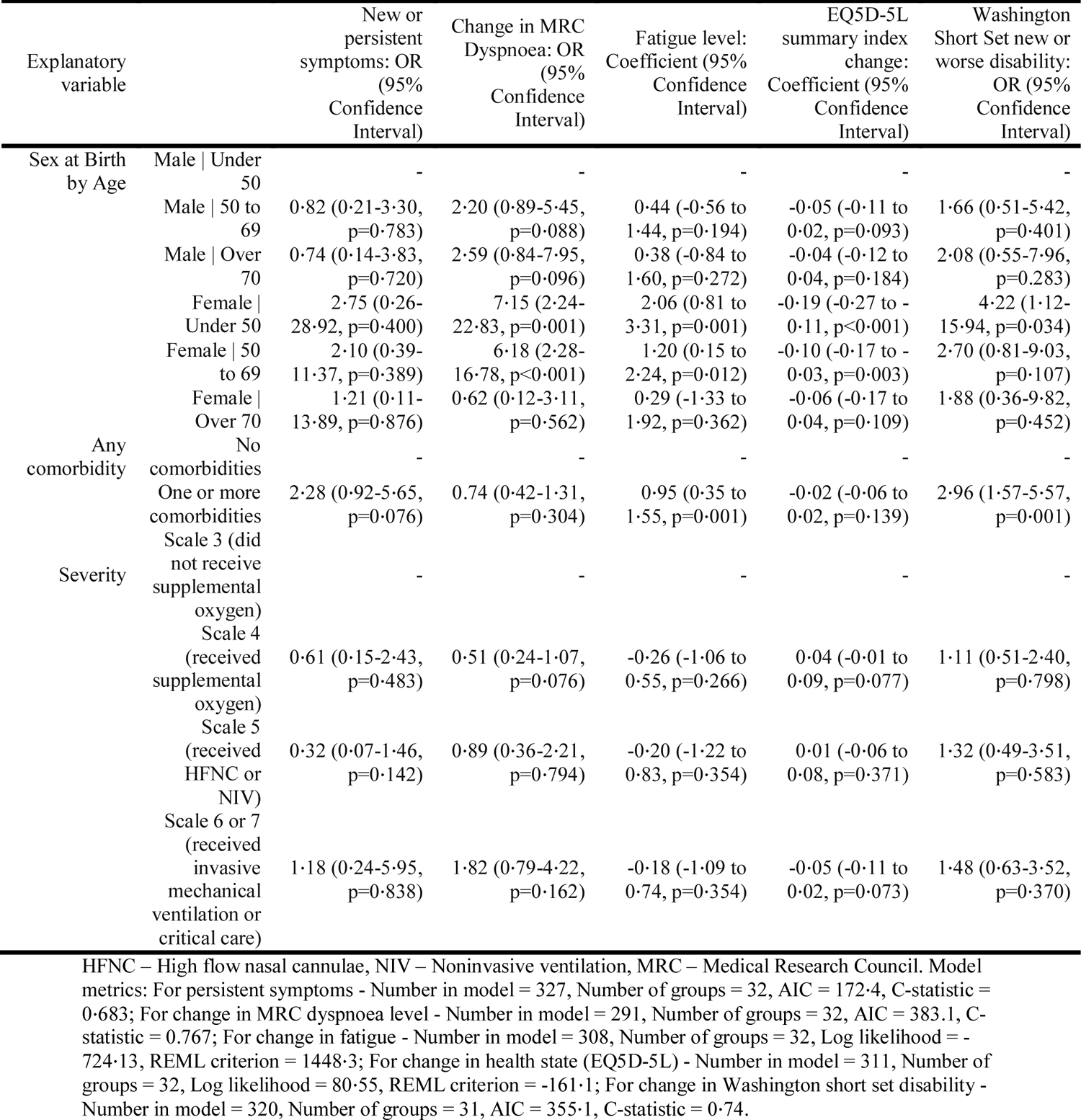
Multilevel regression models for secondary outcomes of new or persistent symptoms, change in MRC dyspnoea scale, fatigue, EQ5D-5L summary index change and Washington Short Set new or worse disability.

Overall summary EQ5D-5L index was 10% lower overall following Covid-19 (median difference - 0 1 points, −0 2 25^th^ centile to 0 0 75^th^ centile, table 2). This change was independent of age or comorbidities (figure 3). The EQ5D-5L dimensions for which most participants reported worsening were usual activities (38 9%, 121/311), anxiety/depression (37 6%, 117/311), and pain/discomfort (37 6%, 117/311) (supplementary table 5). Female sex was significantly associated with increased problems in the usual activity, pain or discomfort and anxiety and depression domains (supplementary table 6).

### Predictors of long-term Covid-19 outcomes

Using multilevel regression models, we adjusted for the effects of age by sex (as this was identified as a significant interaction and retained in our models), the presence of comorbidity and initial in-hospital severity of Covid-19. This generated 6 groups; Males under 50 (34/327), males between 50 and 69 (114/327), males 70 and over (44/327), females under 50 (36/327), females between 50 and 69 (81/327), and females 70 and over (18/327). For the primary outcome of self-reported overall recovery, females under 50 were 5 times less likely to feel fully recovered (figure 4). Similarly, those who received invasive mechanical ventilation were 3 6 times less likely to feel fully recovered (figure 4). For the secondary outcomes, age did not appear to be associated with better or worse long-term outcomes (table 3). Females under 50 were more likely than men to experience persistent fatigue and seven times more likely to experience greater breathlessness, twice as likely to develop new disability and had a significantly poorer health state (EQ5D-5L), all of which persisted in adjusted analyses (table 3). Participants with one or more comorbidities were more likely to experience greater fatigue, disability, and a poorer health state (EQ5D-5L, table 3).

**Figure 5.**
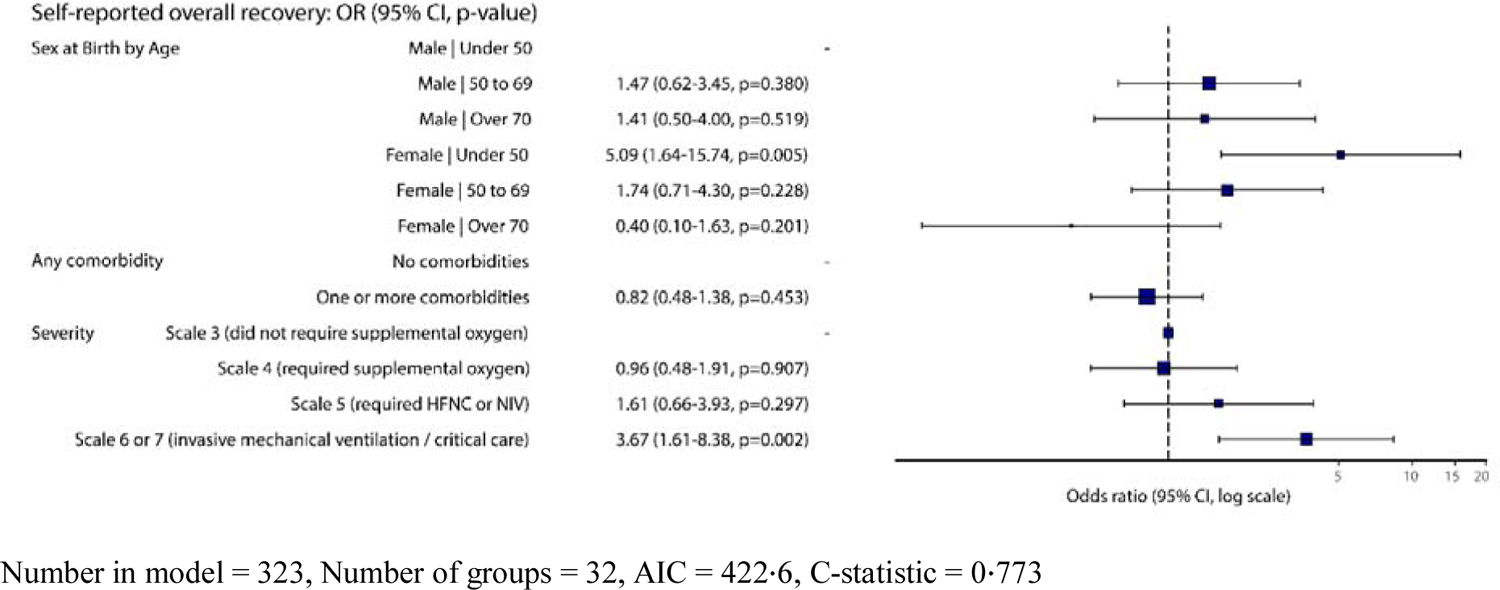
Multilevel model for primary outcome of self-reported recovery (reference level is feeling fully recovered).

## Discussion

We found high rates of long-term symptoms and poor long-term outcomes, which were present several months after hospitalisation for Covid-19. Women under 50, and those with severe acute disease requiring critical care had the worst long-term outcomes even after adjusting for severity of the initial illness. Interestingly, our findings were largely unaffected by existing patient comorbidities or disability.

Our findings add considerably to the current literature, as we identify the main risk factor for worse long-term outcomes are being female and under the age of 50. We also have been able to quantify the significant deterioration in disability and breathlessness-related disability in detail. Many of our findings are largely in agreement with other recent studies in Chinese and Russian populations, which also found high rates of breathlessness and fatigue^4,5^. In the community setting, a recent mobile application-based study, described very high rates of breathlessness (71%) and fatigue (98%) in those reporting symptoms persisting over 28 days ^2^. Interestingly, in our population, the presence of symptoms many months after initial infection are higher than the 76% reported by Huang et al. and three times higher than that reported by Munblit et al. There are several reasons why we have found higher rates, which could be related to those responding to each study, or the severity of disease across the different study populations. The Huang et al. and Munblit et al. studies included very small numbers of patients requiring critical care or mechanical ventilation (1% in Huang et al. and under 2 6% in Munblit et al, in contrast to 28 1% 92/327 in our study), suggesting there are significant differences between these populations and our study population. There may several reasons for this, such as challenges to recruitment of critically unwell patients, differing in-hospital mortality rates, preexisting population comorbidities or pressure on the healthcare systems during the pandemic. Based on data from several countries, the higher rates of participants requiring critical care in our study suggests our data is likely to be more generalisable^30–33^.

In our study, being young, female and having a high severity of acute disease were the strongest independent predictors of poor long-term outcomes. It is unclear why females had the worst outcomes. This could be to do with the effects of initial exposure, where females are more likely to be in industries where exposure to SARS-CoV-2 may be higher ^34^, however recent data suggests teachers do not have greater exposure than other working-age populations and there is emerging evidence of divergent host responses to SARS-CoV-2 infection ^35,36^. Another explanation is that females are more likely to survive severe acute disease than men, so could have worse long-term outcomes as a result. However, in our data, we could not find any differences by sex across several measures of disease severity. A further possibility is that men felt less able or inclined to disclose symptoms. From our findings it is clear more research is required into why females have worse long-term outcomes, particularly as sectors where females are likely to have greater exposure to SARS-CoV-2 are beginning to reopen (e.g. education, hospitality and healthcare).

There are several limitations to our study. First, we were not able to follow all the cases that were discharged from hospital, either because they did not give permission or because they did not respond to repeated requests for information. We attempted to reach non-responders to the survey via telephone follow-up to limit potential for selection bias, but not all could be reached. It is possible that those who did not respond might have been well and therefore uninterested in responding, but it could also be that some were too unwell to respond, had died or moved away. Our results may therefore not be fully representative of with the entire population of those hospitalised with Covid-19. Secondly, we did not include patients hospitalised with other non-Covid-19 illness or a contemporaneous control group, therefore it is unknown if the changes in our outcomes e.g. quality of life, are specific to recovery from Covid-19 or may be linked to other aspects of life during the pandemic. Thirdly, patients only completed the survey at one timepoint, limiting comparison across repeat measures. This also meant retrospective measures asking patients to rate outcomes before their Covid-19 illness were included, which are open to recall bias. Finally, as our study focussed on hospitalised patients primarily from the first wave of infection in the UK, our data cannot be generalised to those with disease managed in the community who comprise the majority of individuals affected by Covid-19.

Future research should focus on establishing the optimal care of this cohort, identifying interventions to test in randomised trials and to identify the mechanisms underlying adverse long-term outcomes. The PHOSP-Covid (Post-HOSPitalisation Covid-19) study is ongoing and will inform patient care by adding to our data on the long-term sequelae of Covid-19, looking at the impact on these of acute and post-discharge interventions, and exploring possible mechanisms ^37^.

## Conclusion

In our study of 327 patients who were discharged alive from hospital, we found most participants reported symptoms months after acute Covid-19 infection. The most common symptoms were fatigue and breathlessness. Participants reported significant difficulties, including increased breathlessness, new or worsened disability and worse quality of life following Covid-19. These symptoms were largely independent of age and prior comorbidity, suggesting that the long-term effects of Covid-19 are determined by factors that differ from those that predict increased mortality. Moreover, the high frequency and severity of long-term symptoms emphasise the importance of long-Covid symptoms and the potential long-term impact on population health and wellbeing.

## Data Availability

Data are available for reuse through a secure data sharing platform. Access is welcome through the ISARIC Independent Data and Material Access Committee (https://isaric4c.net).

## Acknowledgements

This work uses data provided by patients and collected by the NHS as part of their care and support #DataSavesLives. We are extremely grateful to the 2,648 frontline NHS clinical and research staff and volunteer medical students, who collected this data in challenging circumstances; and the generosity of the participants and their families for their individual contributions in these difficult times. In particular, the ISARIC Global Covid-19 Follow up working group and the ISARIC global support centre. We would like to thank Sarah Moore, Romans Matulevics, Liliana Resende, James Lee, Tova Strong and Anneli Sandstrom for administrative, data management, graphic design, formatting and dissemination support. We also acknowledge the support of Janet Diaz, Jeremy J Farrar and Nahoko Shindo.

## ^†^ISARIC Global Covid-19 follow up working group

Adam Ali, John H Amuasi, Andrea Angheben, Valeria Balan. Ibrahim Richard Bangura, Anna Beltrame, Frank Bloos, Lucille Blumberg, Fernando Bozza, Danilo Buonsenso, Gail Carson, Daniel Cassaglia, Muge Cevik, Allegra Chatterjee, Andrew Dagens, Emmanuelle Denis, Yash Doshi, Thomas M. Drake, Murray Dryden, Anne Margarita Dyrhol Riise, Michael Edelstein, Natalie Elkheir, Rob Fowler, Kyle Gomez, Katrina Hann, Ewen M Harrison, Madiha Hashmi, Lars Hegelund, Aquiles Henriquez Trujillo, Antonia Ho, Jan Cato Holter, Jane Ireson, Nina Jamieson, Waasila Jassat, Edwin Jesudason, Anders Benjamin Kildal, Sulaiman Lakoh, Nicola Latronico, James Lee, Wei Shen Lim, Sam Lissaeur, Nazir Lone, David J Lowe, Sinnadurai Manohan, Romans Matulevics, Joanne McPeake, Laura Merson, Roberta Meta, Melina Michelin, Sarah Moore, Ben Morton, Caroline Mudara, Daniel Munblit, Srinivas Murthy, Behzad Nadjm, Piero Olliaro, Carlo Palmieri, Prasan K Panda, Simone Piva, Daniel R Plotkin, Matteo Puntini, Jordi Rello, Liliana Resende, Luis Felipe Reyes, Ishmeala Rigby, Sergio Ruiz Saltana, Clark D Russell, Steffi Ryckaert, Janet T Scott, Malcolm G. Semple, Louise Sigfrid, Girish Sindhwani Pulm, Arne Søraas, Renaud Tamisier, Lance Turtle, Natalie Wright, John Kenneth Baille, Caterina Caminiti, Nikita Nekliudov, Ebrahim Ndure, Caroline Vika

## ^††^ISARIC4C investigators

Consortium lead investigator: J Kenneth Baillie; chief investigator: Malcolm G Semple; co-lead investigator: Peter JM Openshaw; ISARIC clinical coordinator: Gail Carson; *Co-Investigator*: Benjamin Bach, Wendy S Barclay, Debby Bogaert, Meera Chand, Graham S Cooke, Annemarie B Docherty, Jake Dunning, Ana da Silva Filipe, Tom Fletcher, Christoper A Green, Ewen M Harrison, Julian A Hiscox, Antonia Ying Wai Ho, Peter W Horby, Samreen Ijaz, Saye Khoo, Paul Klenerman, Andrew Law, Wei Shen Lim, Alexander J Mentzer, Laura Merson, Alison M Meynert, Mahdad Noursadeghi, Shona C Moore, Massimo Palmarini, William A Paxton, Georgios Pollakis, Nicholas Price, Andrew Rambaut, David L Robertson, Clark D Russell, Vanessa Sancho-Shimizu, Janet T Scott, Thushan de Silva, Louise Sigfrid, Tom Solomon, Shiranee Sriskandan, David Stuart, Charlotte Summers, Richard S Tedder, Emma C Thomson, AA Roger Thompson, Ryan S Thwaites, Lance CW Turtle, Rishi K Gupta, Carlo Palmieri, Maria Zambon, Chloe Donohue, Ruth Lyons, Fiona Griffiths, Wilna Oosthuyzen, Riinu Pius, Thomas M Drake, Cameron J Fairfield, Stephen R Knight, Kenneth A Mclean, Derek Murphy, Catherine A Shaw, Michelle Girvan, Egle Saviciute, Stephanie Roberts, Janet Harrison, Laura Marsh, Marie Connor, Sophie Halpin, Clare Jackson, Carrol Gamble, Andrew Law, Murray Wham, Sara Clohisey, Ross Hendry, James Scott-Brown, Victoria Shaw, Sarah E McDonald, Jane A Armstrong, Milton Ashworth, Innocent G Asiimwe, Siddharth Bakshi, Samantha L Barlow, Laura Booth, Benjamin Brennan, Katie Bullock, Benjamin WA Catterall, Jordan J Clark, Emily A Clarke, Sarah Cole, Louise Cooper, Helen Cox, Christopher Davis, Oslem Dincarslan, Chris Dunn, Philip Dyer, Angela Elliott, Anthony Evans, Lorna Finch, Lewis WS Fisher, Terry Foster, Isabel Garcia-Dorival, William Greenhalf, Philip Gunning, Catherine Hartley, Rebecca L Jensen, Christopher B Jones, Trevor R Jones, Shadia Khandaker, Katharine King, Robyn T. Kiy, Chrysa Koukorava, Annette Lake, Suzannah Lant, Diane Latawiec, Lara Lavelle-Langham, Daniella Lefteri, Lauren Lett, Lucia A Livoti, Maria Mancini, Sarah McDonald, Laurence McEvoy, John McLauchlan, Soeren Metelmann, Nahida S Miah, Joanna Middleton, Joyce Mitchell, Shona C Moore, Ellen G Murphy, Rebekah Penrice-Randal, Jack Pilgrim, Tessa Prince, Will Reynolds, P. Matthew Ridley, Debby Sales, Victoria E Shaw, Rebecca K Shears, Benjamin Small, Krishanthi S Subramaniam, Agnieska Szemiel, Aislynn Taggart, Jolanta Tanianis-Hughes, Jordan Thomas, Erwan Trochu, Libby van Tonder, Eve Wilcock, J. Eunice Zhang, Lisa Flaherty, Nicole Maziere, Emily Cass, Alejandra Doce Carracedo, Nicola Carlucci, Anthony Holmes, Hannah Massey, Nicola Wrobel, Sarah McCafferty, Kirstie Morrice, Alan MacLean, Daniel Agranoff, Ken Agwuh, Dhiraj Ail, Erin L. Aldera, Ana Alegria, Brian Angus, Abdul Ashish, Dougal Atkinson, Shahedal Bari, Gavin Barlow, Stella Barnass, Nicholas Barrett, Christopher Bassford, Sneha Basude, David Baxter, Michael Beadsworth, Jolanta Bernatoniene, John Berridge, Nicola Best, Pieter Bothma, David Chadwick, Robin Brittain-Long, Naomi Bulteel, Tom Burden, Andrew Burtenshaw, Vikki Caruth, David Chadwick, Duncan Chambler, Nigel Chee, Jenny Child, Srikanth Chukkambotla, Tom Clark, Paul Collini, Catherine Cosgrove, Jason Cupitt, Maria-Teresa Cutino-Moguel, Paul Dark, Chris Dawson, Samir Dervisevic, Phil Donnison, Sam Douthwaite, Ingrid DuRand, Ahilanadan Dushianthan, Tristan Dyer, Cariad Evans, Chi Eziefula, Chrisopher Fegan, Adam Finn, Duncan Fullerton, Sanjeev Garg, Sanjeev Garg, Atul Garg, Effrossyni Gkrania-Klotsas, Jo Godden, Arthur Goldsmith, Clive Graham, Elaine Hardy, Stuart Hartshorn, Daniel Harvey, Peter Havalda, Daniel B Hawcutt, Maria Hobrok, Luke Hodgson, Anil Hormis, Michael Jacobs, Susan Jain, Paul Jennings, Agilan Kaliappan, Vidya Kasipandian, Stephen Kegg, Michael Kelsey, Jason Kendall, Caroline Kerrison, Ian Kerslake, Oliver Koch, Gouri Koduri, George Koshy, Shondipon Laha, Steven Laird, Susan Larkin, Tamas Leiner, Patrick Lillie, James Limb, Vanessa Linnett, Jeff Little, Mark Lyttle, Michael MacMahon, Emily MacNaughton, Ravish Mankregod, Huw Masson, Elijah Matovu, Katherine McCullough, Ruth McEwen, Manjula Meda, Gary Mills, Jane Minton, Mariyam Mirfenderesky, Kavya Mohandas, Quen Mok, James Moon, Elinoor Moore, Patrick Morgan, Craig Morris, Katherine Mortimore, Samuel Moses, Mbiye Mpenge, Rohinton Mulla, Michael Murphy, Megan Nagel, Thapas Nagarajan, Mark Nelson, Matthew K. O’Shea, Igor Otahal, Marlies Ostermann, Mark Pais, Selva Panchatsharam, Danai Papakonstantinou, Hassan Paraiso, Brij Patel, Natalie Pattison, Justin Pepperell, Mark Peters, Mandeep Phull, Stefania Pintus, Jagtur Singh Pooni, Frank Post, David Price, Rachel Prout, Nikolas Rae, Henrik Reschreiter, Tim Reynolds, Neil Richardson, Mark Roberts, Devender Roberts, Alistair Rose, Guy Rousseau, Brendan Ryan, Taranprit Saluja, Aarti Shah, Prad Shanmuga, Anil Sharma, Anna Shawcross, Jeremy Sizer, Manu Shankar-Hari, Richard Smith, Catherine Snelson, Nick Spittle, Nikki Staines, Tom Stambach, Richard Stewart, Pradeep Subudhi, Tamas Szakmany, Kate Tatham, Jo Thomas, Chris Thompson, Robert Thompson, Ascanio Tridente, Darell Tupper-Carey, Mary Twagira, Andrew Ustianowski, Nick Vallotton, Lisa Vincent-Smith, Shico Visuvanathan, Alan Vuylsteke, Sam Waddy, Rachel Wake, Andrew Walden, Ingeborg Welters, Tony Whitehouse, Paul Whittaker, Ashley Whittington, Padmasayee Papineni, Meme Wijesinghe, Martin Williams, Lawrence Wilson, Sarah Cole, Stephen Winchester, Martin Wiselka, Adam Wolverson, Daniel G Wooton, Andrew Workman, Bryan Yates, Peter Young.

## Declarations and statements

### Contributors

JTS, LS, MGS developed the concept of the follow up study. JTS, LS, LWS, TMD, ECJ, WLS, CB, DJL, MC, JMcP, NL, EMH, DM, CDR, AH, LT, and the ISARIC Global Covid-19 follow up working group developed the follow up protocol and methodology. ISARIC4C investigators recruited patients into the study during the admission and entered acute phase data. PB, CD, HH, RS, JH, AG, LM coordinated follow up survey distribution, data entry, AG, AC, LG conducted telephone follow up. LS, JTS, GC, HH coordinated resources. LS, JTS, TMD, EP, AD, PO, EH, ABD were involved in data visualisation. TMD, EP, LS, JTS, MEO’H, CH, CEH, PJMO, JKB, CH, ABD, PO, MGS analysed and interpreted the data. LS, TMD, EP, JTS wrote the original draft of the manuscript. All authors reviewed, and revised the manuscript prior to submission. JTS is the guarantor.

## Competing interests

### Ethical approval

Ethical approval was given by the South Central - Oxford C Research Ethics Committee in England (ref 13/SC/0149) and the Scotland A Research Ethics Committee (ref 20/SS/0028).

### Transparency

The lead author (the manuscript’s guarantor) affirms that the manuscript is an honest, accurate, and transparent account of the study being reported; that no important aspects of the study have been omitted; and that any discrepancies from the study as planned (and, if relevant, registered) have been explained.

### Dissemination plans

Dissemination to participants and related patient and public communities: ISARIC4C has a public facing website [ISARIC4C.net] and twitter account (<u>@CCPUKstudy</u>). We are engaging with print and internet press, television, radio, news, and documentary programme makers.

## Supplementary appendix 1

**Supplementary Figure 1.**
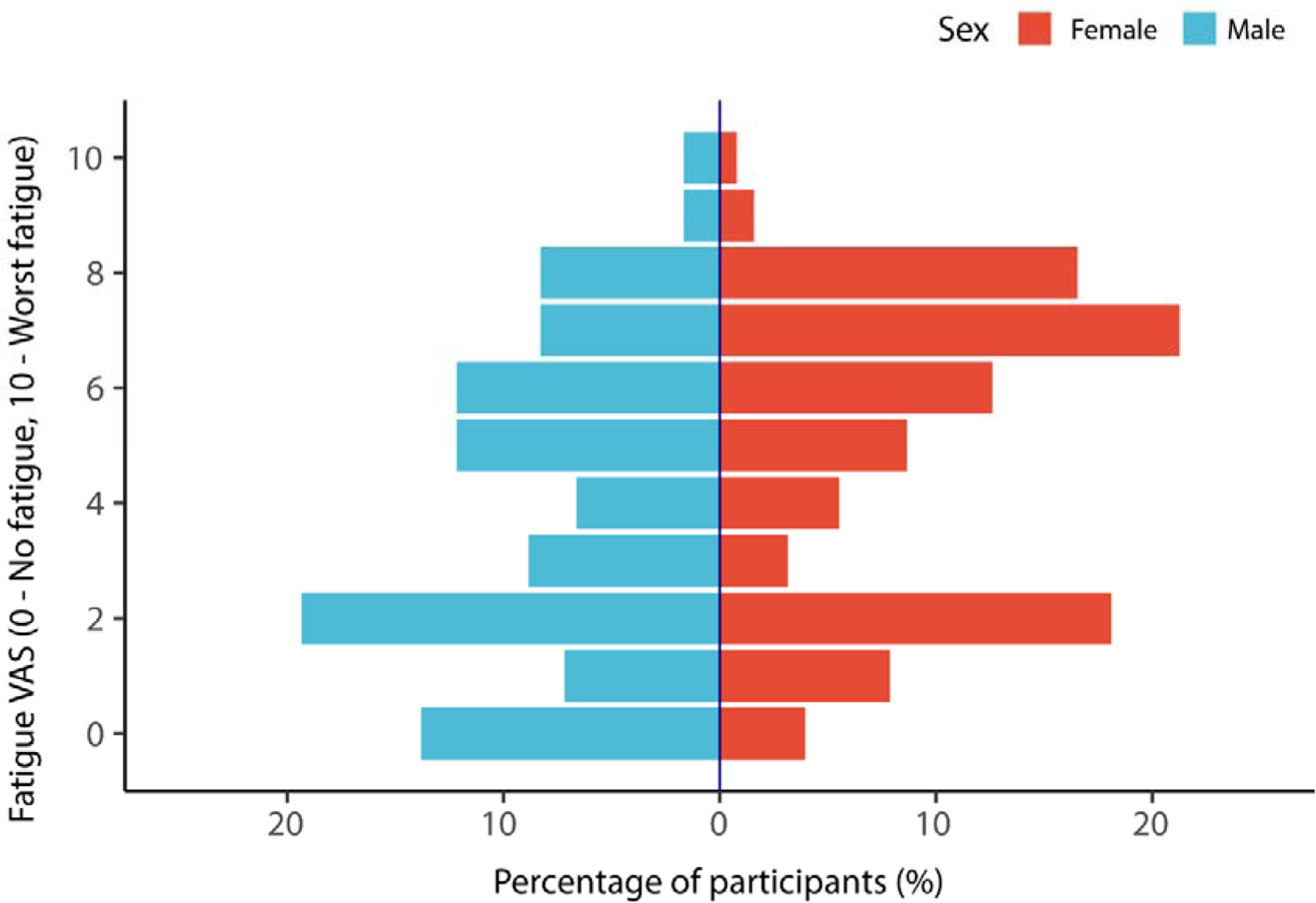
Fatigue rating on Visual Analogue Scale (VAS) by sex.

**Supplementary table 1.**
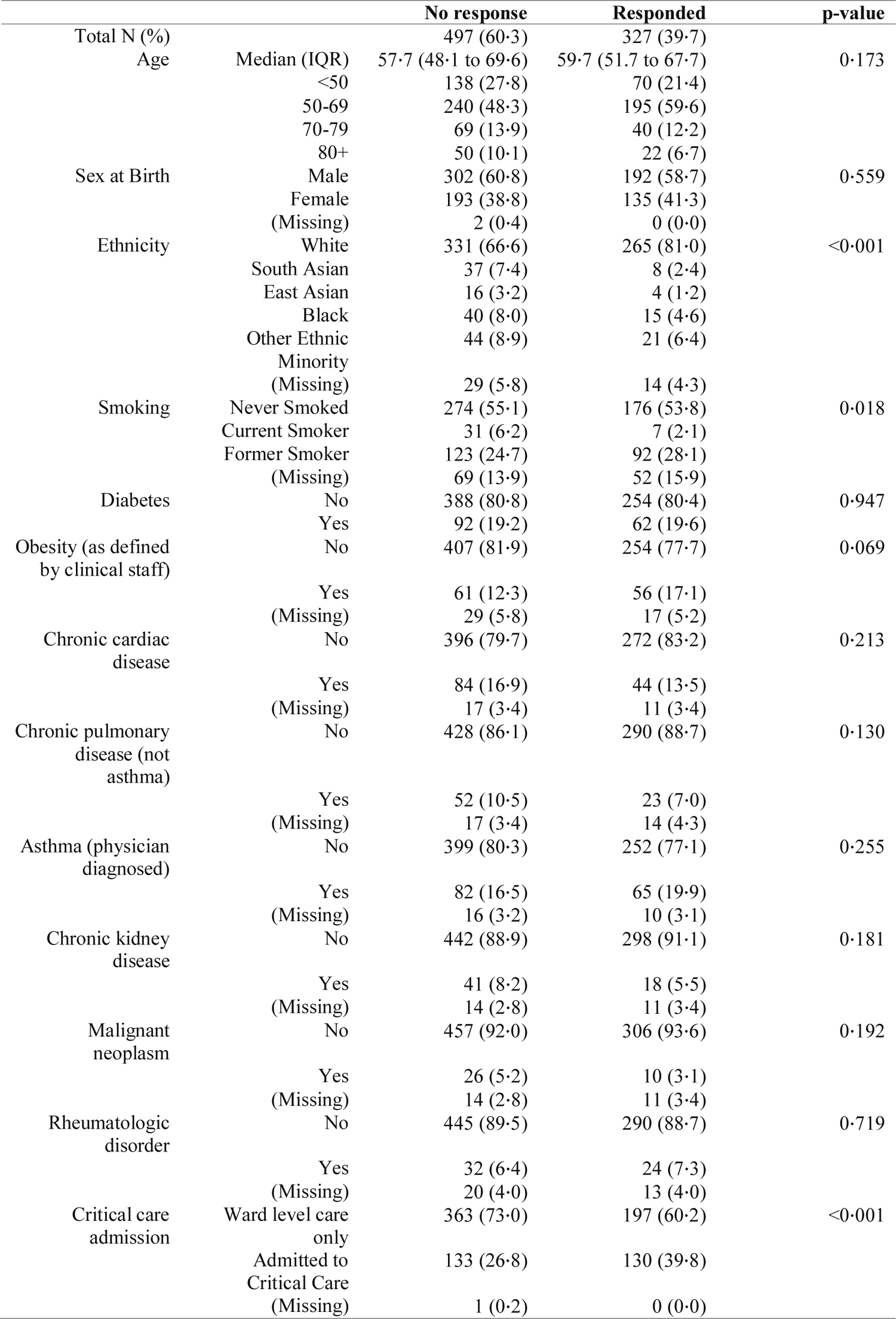
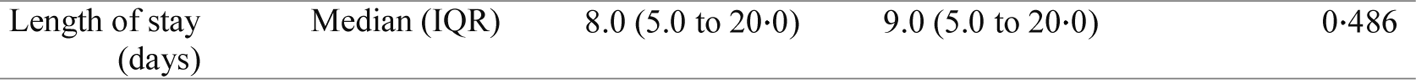
Comparison between respondents and those who did not respond

**Supplementary table 2.** Long-term outcomes by sex

|  |  | Male | Female | p-value |
| --- | --- | --- | --- | --- |
| Total N (%) |  | 192 (58.7) | 135 (41.3) |  |
| Self-reported overall recovery | Feels fully recovered | 56 (29.2) | 27 (20.0) | 0.117 |
|  | Does not feel fully recovered | 103 (53.6) | 76 (56.3) |  |
|  | Not sure | 31 (16.1) | 30 (22.2) |  |
|  | (Missing) | 2 (1.0) | 2 (1.5) |  |
| New or persistent symptoms | No new or persistent symptoms | 17 (8.9) | 5 (3.7) | 0.108 |
|  | New or persistent symptoms | 175 (91.1) | 130 (96.3) |  |
| Change in breathlessness after COVID-19 (MRC Dyspnoea) | No change | 88 (45.8) | 39 (28.9) | 0.015 |
|  | Less breathless | 7 (3.6) | 4 (3.0) |  |
|  | More breathless | 80 (41.7) | 73 (54.1) |  |
|  | (Missing) | 17 (8.9) | 19 (14.1) |  |
| Fatigue level (0 to 10 VAS) | Median (IQR) | 4.0 (2.0 to 6.0) | 6.0 (2.0 to 7.0) | <0.001 |
| EQ5D-5L change in overall summary index | Median (IQR) | -0.0 (-0.2 to 0.0) | -0.1 (-0.3 to 0.0) | <0.001 |
| Washington Group Short Set | No change in disability | 150 (78.1) | 91 (6.4) | 0.059 |
|  | New or worse disability in at least one domain | 39 (20.3) | 40 (29.6) |  |
|  | (Missing) | 3 (1.6) | 4 (3.0) |  |

HFNC – High flow nasal cannulae, NIV – Noninvasive ventilation, MRC – Medical Research Council, IQR – Interquartile range, presented as 25<sup>th</sup> to 75<sup>th</sup> centiles. Numbers are presented as N (%), unless otherwise denoted as a continuous variable.

**Supplementary table 3.**
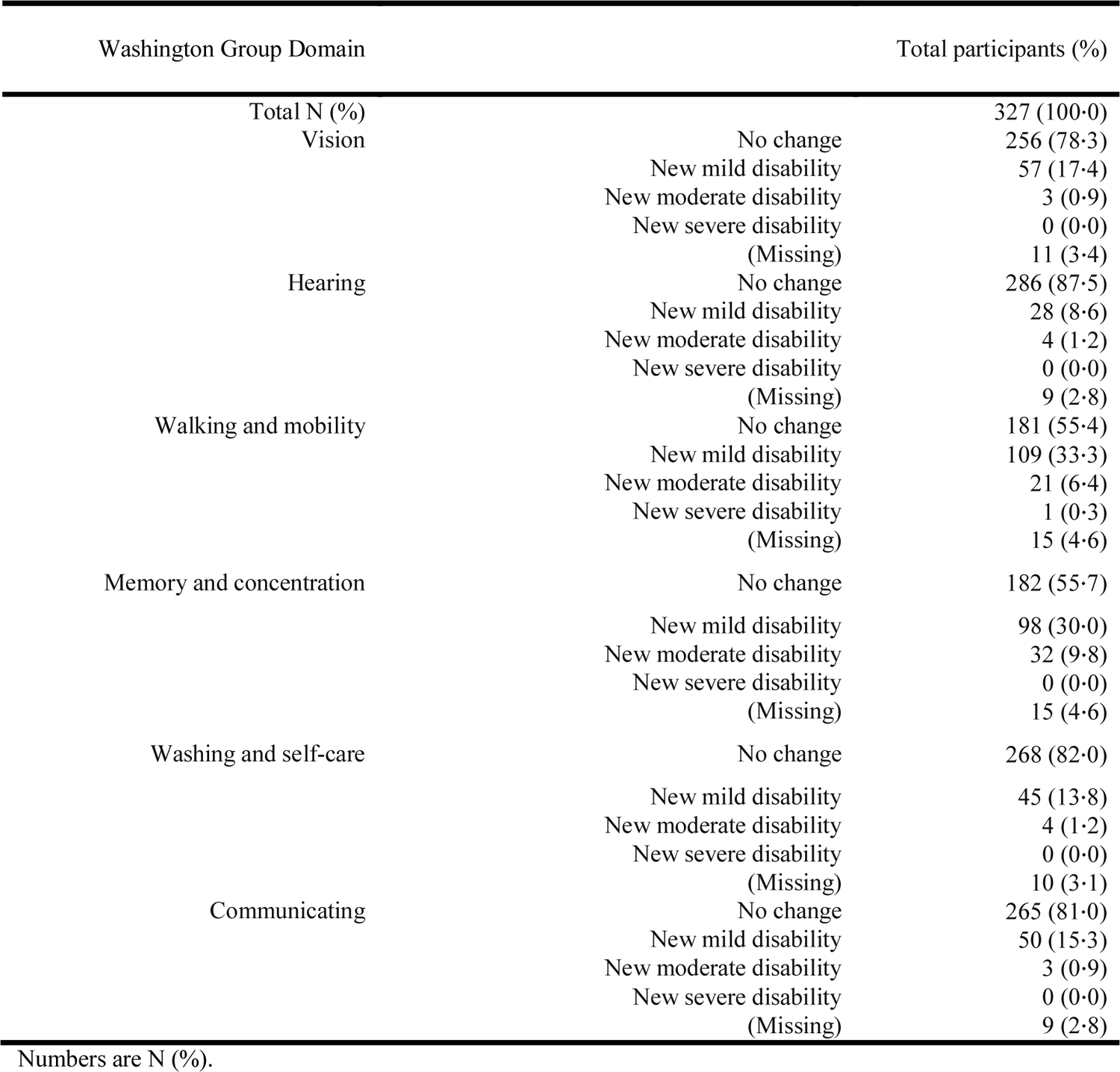
Overall new or worse disability by Washington Group disability domains before onset of Covid-19 compared with disability at time of follow-up.

**Supplementary table 4.** New or worse disability across Washington Group disability domains, before onset of Covid-19 compared with disability at time of follow-up stratified by sex.

| Washington Group Domain |  | Male | Female | p-value |
| --- | --- | --- | --- | --- |
| Total N (%) |  | 192 (58.7) | 135 (41.3) |  |
| Vision | No change | 160 (83.3) | 96 (71.1) | 0.009 |
|  | New mild disability | 23 (12.0) | 34 (25.2) |  |
|  | New moderate disability | 2 (1.0) | 1 (0.7) |  |
|  | New severe disability | 0 (0.0) | 0 (0.0) |  |
|  | (Missing) | 7 (3.6) | 4 (3.0) |  |
| Hearing | No change | 168 (87.5) | 118 (87.4) | 0.246 |
|  | New mild disability | 17 (8.9) | 11 (8.1) |  |
|  | New moderate disability | 4 (2.1) | 0 (0.0) |  |
|  | New severe disability | 0 (0.0) | 0 (0.0) |  |
|  | (Missing) | 3 (1.6) | 6 (4.4) |  |
| Walking and mobility | No change | 120 (62.5) | 61 (45.2) | 0.040 |
|  | New mild disability | 55 (28.6) | 54 (40.0) |  |
|  | New moderate disability | 11 (5.7) | 10 (7.4) |  |
|  | New severe disability | 1 (0.5) | 0 (0.0) |  |
|  | (Missing) | 5 (2.6) | 10 (7.4) |  |
| Memory and concentration | No change | 119 (62.0) | 63 (46.7) | 0.023 |
|  | New mild disability | 52 (27.1) | 46 (34.1) |  |
|  | New moderate disability | 14 (7.3) | 18 (13.3) |  |
|  | New severe disability | 0 (0.0) | 0 (0.0) |  |
|  | (Missing) | 7 (3.6) | 8 (5.9) |  |
| Washing and self-care | No change | 161 (83.9) | 107 (79.3) | 0.651 |
|  | New mild disability | 24 (12.5) | 21 (15.6) |  |
|  | New moderate disability | 2 (1.0) | 2 (1.5) |  |
|  | New severe disability | 0 (0.0) | 0 (0.0) |  |
|  | (Missing) | 5 (2.6) | 5 (3.7) |  |
| Communicating | No change | 159 (82.8) | 106 (78.5) | 0.552 |
|  | New mild disability | 26 (13.5) | 24 (17.8) |  |
|  | New moderate disability | 2 (1.0) | 1 (0.7) |  |
|  | New severe disability | 0 (0.0) | 0 (0.0) |  |
|  | (Missing) | 5 (2.6) | 4 (3.0) |  |

Numbers are N (%).

**Supplementary table 5.**
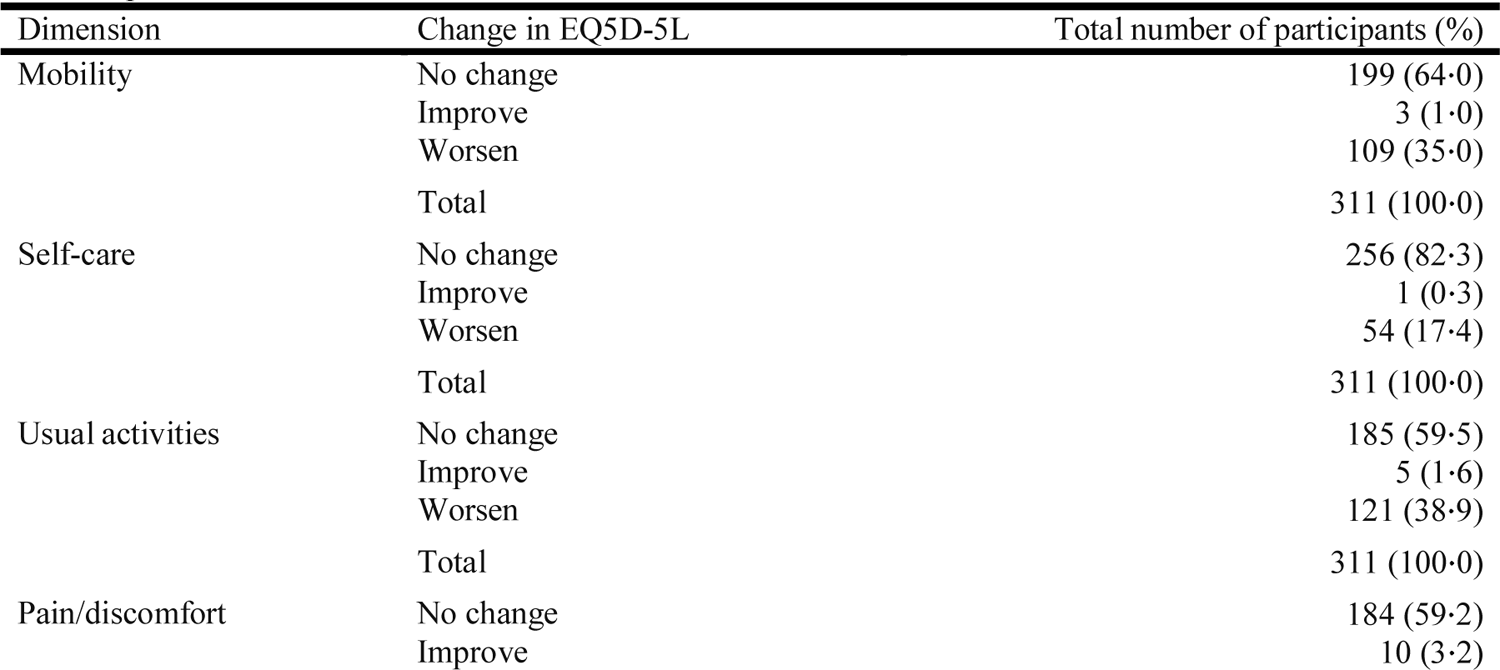
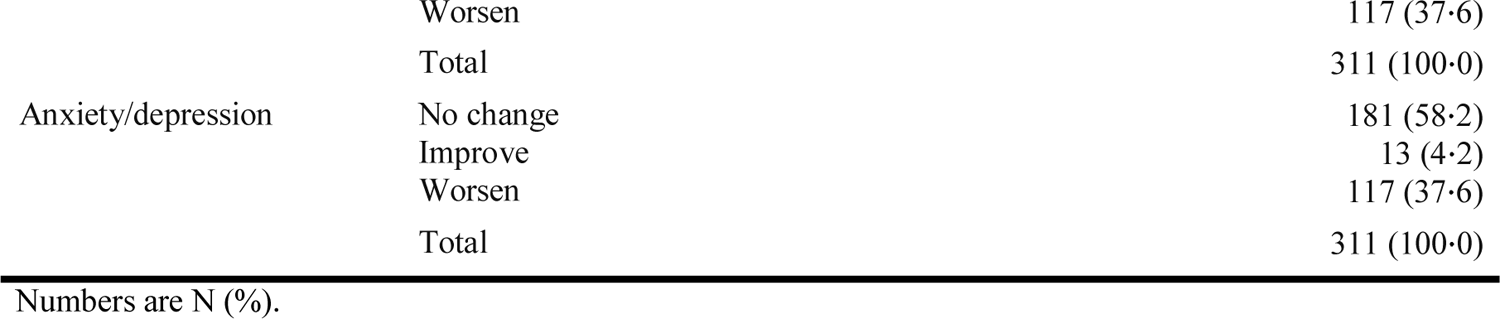
Overall changes by EQ5D-5L dimension before Covid-19 onset and at time of follow-up.

**Supplementary table 6.** Overall changes by EQ5D-5L dimension and sex before Covid-19 onset and at time of follow-up.

| Dimension | Change in EQ5D-5L | Male (%) | Female (%) |
| --- | --- | --- | --- |
| Mobility | No change | 126 (68·1) | 73 (57·9) |
|  | Improve | 2 (1·1) | 1 (0·8) |
|  | Worsen | 57 (30·8) | 52 (41·3) |
|  | Total | 185 (100·0) | 126 (100·0) |
| Self-care | No change | 156 (84·3) | 100 (79·4) |
|  | Improve | 1 (0·5) | 0 (0·0) |
|  | Worsen | 28 (15·1) | 26 (20·6) |
|  | Total | 185 (100·0) | 126 (100·0) |
| Usual activities | No change | 120 (64·9) | 65 (51·6) |
|  | Improve | 3 (1·6) | 2 (1·6) |
|  | Worsen | 62 (33·5) | 59 (46·8) |
|  | Total | 185 (100·0) | 126 (100·0) |
| Pain/discomfort | No change | 121 (65·4) | 63 (50·0) |
|  | Improve | 6 (3·2) | 4 (3·2) |
|  | Worsen | 58 (31·4) | 59 (46·8) |
|  | Total | 185 (100·0) | 126 (100·0) |
| Anxiety/depression | No change | 126 (68·1) | 55 (43·7) |
|  | Improve | 8 (4·3) | 5 (4·0) |
|  | Worsen | 51 (27·6) | 66 (52·4) |
|  | Total | 185 (100·0) | 126 (100·0) |

Numbers are N (%).

**Supplementary table 7.** Proportion of participants experiencing new or persistent symptoms at time of follow-up.

| Symptom | Number of participants experiencing symptom (%) | Participant denominator |
| --- | --- | --- |
| Fatigue | 255 (82.8) | 308 |
| Shortness of breath | 175 (53.5) | 327 |
| Problems sleeping | 151 (46.2) | 327 |
| Headache | 129 (39.4) | 327 |
| Limb weakness | 123 (37.6) | 327 |
| Joint pain or swelling | 121 (37.0) | 327 |
| Persistent muscle pain | 121 (37.0) | 327 |
| Dizziness/light headedness | 117 (35.8) | 327 |
| Problems with balance | 99 (30.3) | 327 |
| Swollen ankle | 80 (24.5) | 327 |
| Palpitations | 76 (23.2) | 327 |
| Constipation | 59 (18.0) | 327 |
| Problems seeing | 59 (18.0) | 327 |
| Diarrhoea | 58 (17.7) | 327 |
| Stomach pain | 58 (17.7) | 327 |
| Chest pains | 50 (15.3) | 327 |
| Persistent cough | 50 (15.3) | 327 |
| Erectile dysfunction | 45 (23.4) | 192 |
| Pain on breathing | 43 (13.1) | 327 |
| Loss of smell | 42 (12.8) | 327 |
| Other | 42 (12.8) | 327 |
| Persistent fevers | 36 (11.0) | 327 |
| Loss of taste | 35 (10.7) | 327 |
| Nausea/vomiting | 34 (10.4) | 327 |
| Loss of appetite | 33 (10.1) | 327 |
| Problems swallowing | 28 (8.6) | 327 |
| Skin rash | 27 (8.3) | 327 |
| Weight loss | 24 (7.3) | 327 |
| Problems passing urine | 23 (7.0) | 327 |
| Hemiplegia/paraesthesiae | 20 (6.1) | 327 |
| Toe lesions | 13 (4.0) | 327 |

